# The effect of Covid-19 isolation measures on the cognition and mental health of people living with dementia: a rapid systematic review of one year of evidence

**DOI:** 10.1101/2021.03.17.21253805

**Authors:** Aida Suárez-González, Jayeeta Rajagopalan, Gill Livingston, Suvarna Alladi

**Affiliations:** Dementia Research Centre, UCL Queen Square Institute of Neurology, UCL, London, UK; Strengthening Responses to Dementia in Developing Countries (STRiDE) India, National Institute of Mental Health and Neurosciences, Bangalore, India; Division of Psychiatry, Maple House, University College London; Department of Neurology, National Institute of Mental Health and Neurosciences Bangalore, India

**Author notes:** Correspondence: Aida Suárez-González, UCL Queen Square Institute of Neurology, Dementia Research Centre, box 16, 8-11 Queen Square, London WC1N 3BG.

**Keywords:** dementia, Covid-19, lockdown, isolation, neuropsychiatric symptoms

## Abstract

**Background:** Covid-19 control policies have entailed lockdowns and confinement. Although these isolation measures are thought to be particularly hard and possibly harmful to people with dementia, their specific impact during the pandemic has not yet been synthesised. We aimed to examine and summarise the global research evidence describing the effect of Covid-19 isolation measures on the health of people living with dementia.

**Method:** We searched Pubmed, PsycINFO and CINAHL up to February 2021 for peer-reviewed quantitative studies of the effects of isolation measures during Covid-19 on cognitive, psychological and functional symptoms of people with any kind of dementia or mild cognitive impairment. We summarised the findings of included papers following current guidelines for rapid reviews.

**Results:** We identified 15 eligible papers, examining a total of 6,442 people with dementia. 13/15 were conducted in people living in the community and 2 in care homes. 60% (9/15) studies reported changes in cognition with 77% (7/9) of them describing declined cognition by >50% of respondents. 93% (14/15) of studies reported worsening or new onset of behavioural and psychological symptoms. 46% (7/15) studies reported changes in daily function, 6 of them reporting a functional decline in a variable proportion of the population studied.

**Conclusion:** Lockdowns and confinement measures brought about by the pandemic have damaged the cognitive and psychological health and functional abilities of people with dementia across the world. It is urgent that infection control measures applied to people with dementia are balanced against the principles of non-maleficence. This systematic review makes 4 specific calls for action.

**Key Points:**

- Neuropsychiatric symptoms of people with dementia (e.g., anxiety, depressive symptoms, apathy, agitation) were found to worsen during lockdown in the majority of studies.
- Cognitive decline affecting memory, orientation concentration and communication was observed by caregivers within few weeks after lockdown.
- The deterioration reported occurred in a short window of time (between 1 and 4 months) and it is unlikely to be attributable to the natural variation of the course of dementia.
- There is little research conducted in care home residents with dementia (only 2 papers found).
- Increase consumption of antipsychotics and benzodiazepines has occurred in people with dementia during lockdown.
- Evidence indicates that isolation measures quickly damaged people’s with dementia cognitive and mental health and probably accelerated overall decline.

## 1. Introduction

The first wave of the new coronavirus disease, Covid-19, swept across the world in March 2020 becoming a universal health challenge. In that month, many countries declared national lockdowns or imposed strict restrictions to try to reduce virus transmission. With little knowledge available at the time about the best way to respond to Covid-19, authorities world-wide enforced strict measures to hugely suppress physical contact. Such measures have evolved, coming and going, as new waves of the virus unfolded, with both direct and collateral significance for individuals. People with dementia have great vulnerability to Covid-19 infection and death but also to the indirect consequences of lockdown and confinement as they usually require day-to-day help and may not be able either to understand the rapidly changing situation and associated knowledge or to make informed decisions about responding to it (Liu et al., 2021). The challenge of living with dementia is amplified by the reduction in therapeutic and essential services because of Covid-19 containment measures, which have entailed disruption in services for maintaining health and well-being and the good management of the dementia-related symptoms (Hanna et al., 2020, Giebel et al., 2020). For instance, not only were specialist dementia consultations postponed or suspended but day centres and home services and most therapeutic and support activities were closed or curtailed in many countries (Giebel et al., 2020, Liu et al., 2021). These are vital services for people with dementia in that the focus of dementia treatment and care is on quality of life (Livingston et al., 2020) and maintaining function for as long as possible, regaining lost function when possible, and adapting to lost function that cannot be regained (Poulos et al., 2017). In some countries the restrictions applied during lockdowns barely allowed for one hour a day of outdoor activities while in others not even that was permitted. Even those people with dementia who lived with relatives or partners were more isolated than before. For those residing in care homes it was even harder, since the measures in residential facilities have been considerably more restrictive and prolonged and maintained for the year after the start of the pandemic. To reduce infection and death rates in care and nursing homes residents who have been in contact with possible infection have been isolated for weeks in their own room and many homes have had a total ban on visitors (including children and spouses) (Comas-Herrera et al., 2020).

### Effects of isolation on people with dementia

Since the early months of the pandemic, families, healthcare professionals and scientist have raised concerns about the potentially adverse consequences of confinement and isolation measures on the health of people with dementia who are already vulnerable (Suárez-González, 2020; Suárez-González et al. 2020a). This alarm was because confinement and stimulation deprivation is the opposite of what is therapeutically recommended for someone living with dementia (NICE 2018). People with dementia are susceptible to changes in their routines because these are experienced as stressful events that impose an extra cognitive load on someone who is cognitively impaired. For instance, changes in well-established daily patterns may affect the use of strategies routinely adopted to manage the dementia symptoms and daily living. A lack of activities, sensory stimulation and pleasure make people with dementia more vulnerable to boredom, anxiety, apathy sleep disturbance, agitation, hallucinations and these neuropsychiatric symptoms may lead to conflicts with family. Lastly, while staying at home, time spent in sedentary activities increases and daily physical activity is reduced, which may reduce physical function and independence. These symptoms result from a combination of factors: biological (product of the neuropathological changes of the brain caused by the disease), psychological, environmental and social, as well as unmet needs (responsive behaviours). As the first studies on the topic emerged, those concerns started to crystalise (Canevelli t al., 2020; Lara et al., 2020; Suzuki et al., 2020). The presence of significant NPS is associated with more rapid progression to severe dementia and death (Peters et al., 2015) and negatively impacts on the quality of life of the person with the condition.

Knowledge of the impact of isolation and confinement of people with dementia during the pandemic on their health is crucial to balance the risks and benefits of the current public health policies, the provision of care in post-emergency scenarios and future waves. The purpose of the current study is to bring together the evidence about impact to ensure that no person with dementia is left behind.

This rapid review seeks to answer the following questions: 1) What is the relationship between Covid-19 isolation measures and the cognitive, psychological symptoms functional level among people with dementia living in the community?

2) What is the relationship between Covid-19 isolation measures and the cognitive and psychological symptoms and functional level among people with dementia living in care homes?

## 2. Methods/Design

The protocol for this rapid review was registered in PROSPERO (CRD42021229259) (Suárez-González et al., 2021). We followed PRISMA guidelines (Preferred Reporting Items for Systematic Reviews and Meta-Analyses) for conducting the review and preparing the report (13,14). We systematically reviewed the literature about Covid-19, isolation measures and the cognitive, mental health and functional level of people with dementia using the recommended general framework (Tricco et al., 2017) (Garrity et al., 2021).

### 2.1 Selection of studies

We included studies that described the effects of isolation measures (i.e. lockdowns) on the cognitive, psychological and functional performance of people with any kind of dementia or mild cognitive impairment. Inclusion criteria required studies to report either a percentage of people with dementia showing changes due to isolation for at least one of the three variables of interest (cognition, psychological symptoms of level on function for activities of daily living) or a mean and standard deviation of any of these measures at a group level during the pandemic. The outcome measures were 1) cognitive function and 2) psychological symptoms and 3) level of performance for activities of daily living (functional level).

We excluded studies providing information about the variables of interest during the pandemic without referring to any change due to isolation measures, studies in languages other than English, conference abstracts, grey literature and non-peer-reviewed material (pre-prints were excluded).

### 2.2. Information sources and search terms

We searched the following databases: Pubmed, PsycINFO and CINAHL. The literature included was indexed, published and peer-reviewed literature with unlimited date range up until 27^th^ February 2021. Database specific conventions and the use of multiple search fields was customised for individual databases. To streamline the review process according to frameworks recommended for rapid reviews, reference lists from key articles and reviews were not checked. The following search terms were developed in consultation with experienced researchers and librarians and piloted before being used in the current review: (dementia OR neurocognitive disorders OR cognitive impairment) AND (covid* OR coronavirus* OR coronavirus) AND (pandemic OR outbreak OR lockdown OR confinement OR isolation OR quarantine) AND (behavioral symptoms OR behavioural symptom* OR behavioral symptom* OR psychological symptom* OR psychological well-being OR psychological wellbeing OR loneliness OR loneliness OR neuropsychiatric symptom* OR mental health OR physical health OR mood* OR communication OR communication OR depression OR depressi* OR anxiety OR anxiety OR apathy OR apathy OR agitation OR irritab* OR wandering OR insomnia OR sleep* OR functional decline OR cognitive decline OR activities of daily living OR hallucinations OR hallucinat* OR delusions OR delusion*OR facing OR coping).

The search strategy for the three databases is fully reported in Appendix 1. Results were exported into Endnote ⍰ software version X9 and automatically deduplicated. A multi-level title-first method was conducted to screen and select the candidate articles, screen titles first and abstracts after (Mateen et al., 2013). The main reviewer (ASG) conducted screening and selection and applied inclusion and exclusion criteria. A 10% sample of the full-text articles selected and 10% of the excluded were double screened (JR).

### 2.3. Data extraction and quality assessment

Data extraction from each article was completed by one reviewer, verified by a second one independently and recorded in a standards extraction form covering sample size, measure used, time of data collection, decline in cognition, appearance or worsening of behavioural/psychological symptoms, decline in activities of daily living, increase or addition of pharmacological therapy and quality score. We used the Joanna Briggs Institute (JBI) critical appraisal tools (https://joannabriggs.org/critical-appraisal-tools) for quality assessment to describe the characteristics of the studies included and current body of knowledge on the impact of Covid-19 isolation measures on the cognitive, psychological and functional health of people with dementia. The assessment was done on study design, with quality assessment conducted by a reviewer (ASG) and 3 (20) % of scoring verified by a second one (JR). Discrepancies we resolved between raters.

As recommended in JBI manual for evidence synthesis, we considered as good quality those studies achieving a cut-off score of more than 70% of the items assessed for the JBI critical appraisal tools, which is fewer than eight out of 11 for cohort studies; and fewer than five out of eight for analytical cross-sectional studies ‘‘yes’’ answers (Tufanaru et al., 2017)

### 2.4. Synthesis of results

Studies were grouped according to whether they addressed people with dementia living in the community or in care homes and by study design.

## 3. Results

Figure 1 presents a PRISMA flow diagram of the article inclusion process. The studies’ characteristics are summarised in Table 1 and Table 2 by order of their quality score. Table 1 reports studies conducted on people with dementia living in the community and is divided into those providing a measure pre and post lockdown and those providing a mean measure during lockdown. Table 2 illustrates the studies conducted on people with dementia living in care homes. We identified 15 studies examining the effects of lockdown on a total sample of 6,442 people with dementia. 13 of them were conducted with people living in the community (2 of them collecting data pre- and post-pandemic and 11 cross sectional) and 2 in care homes. All studies conducted the cross-sectional data collection within the first months of lockdown or right after, while two of them show measures collected prospectively at baseline before and then after lockdown (Lara et la., 2020, Borges-Machado et al., 2020).

**Figure 1.**
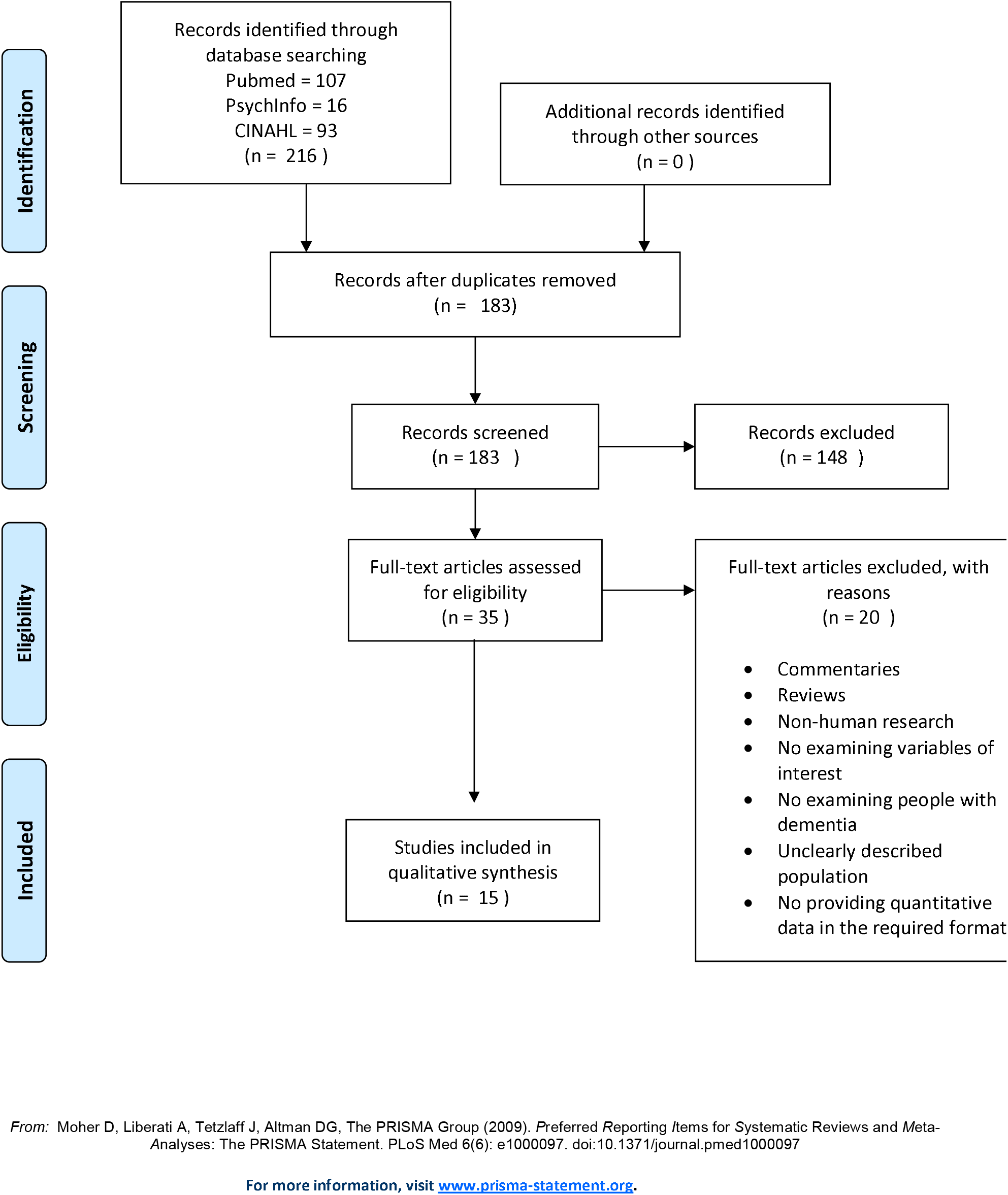
Prisma Flow Diagram.

**Table 1.**
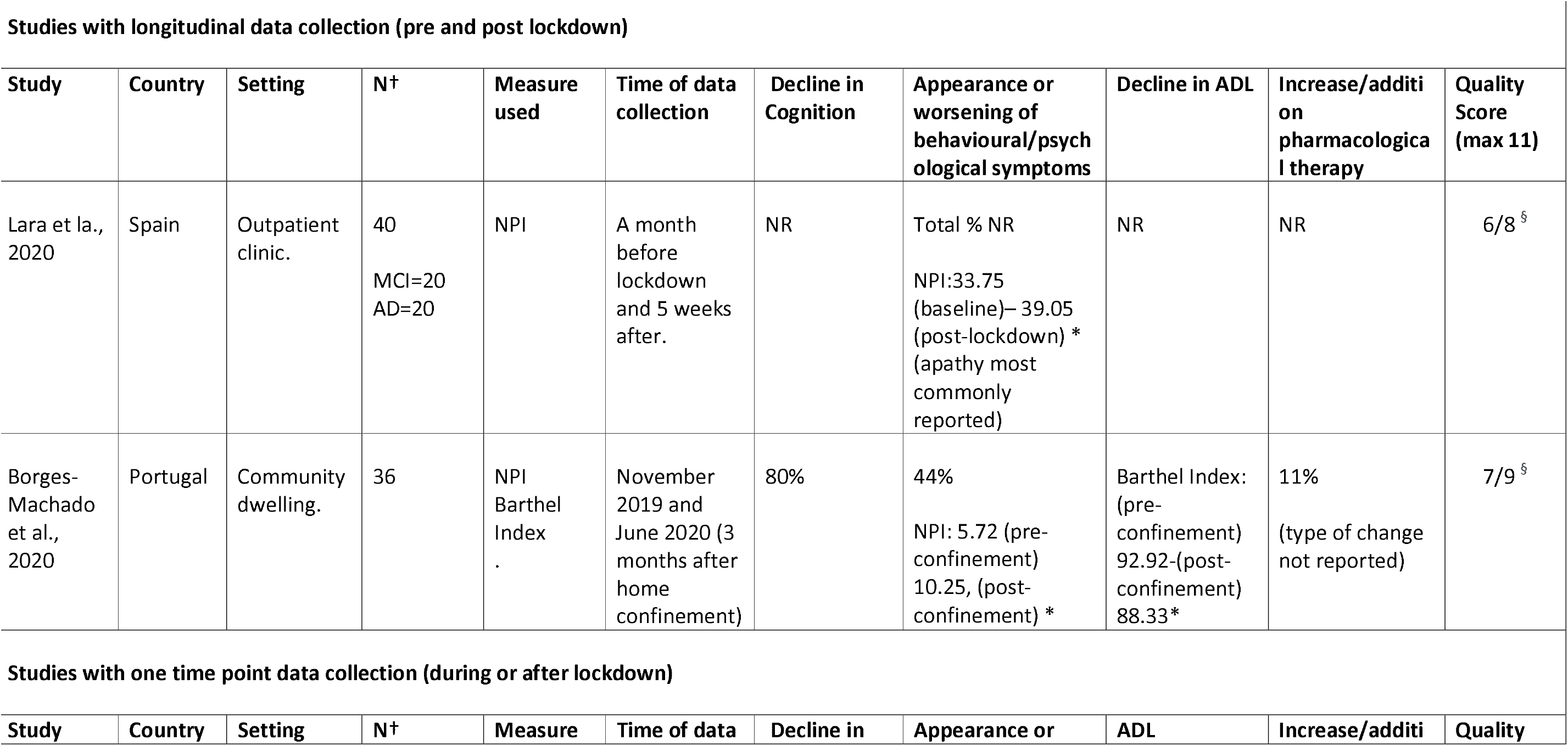
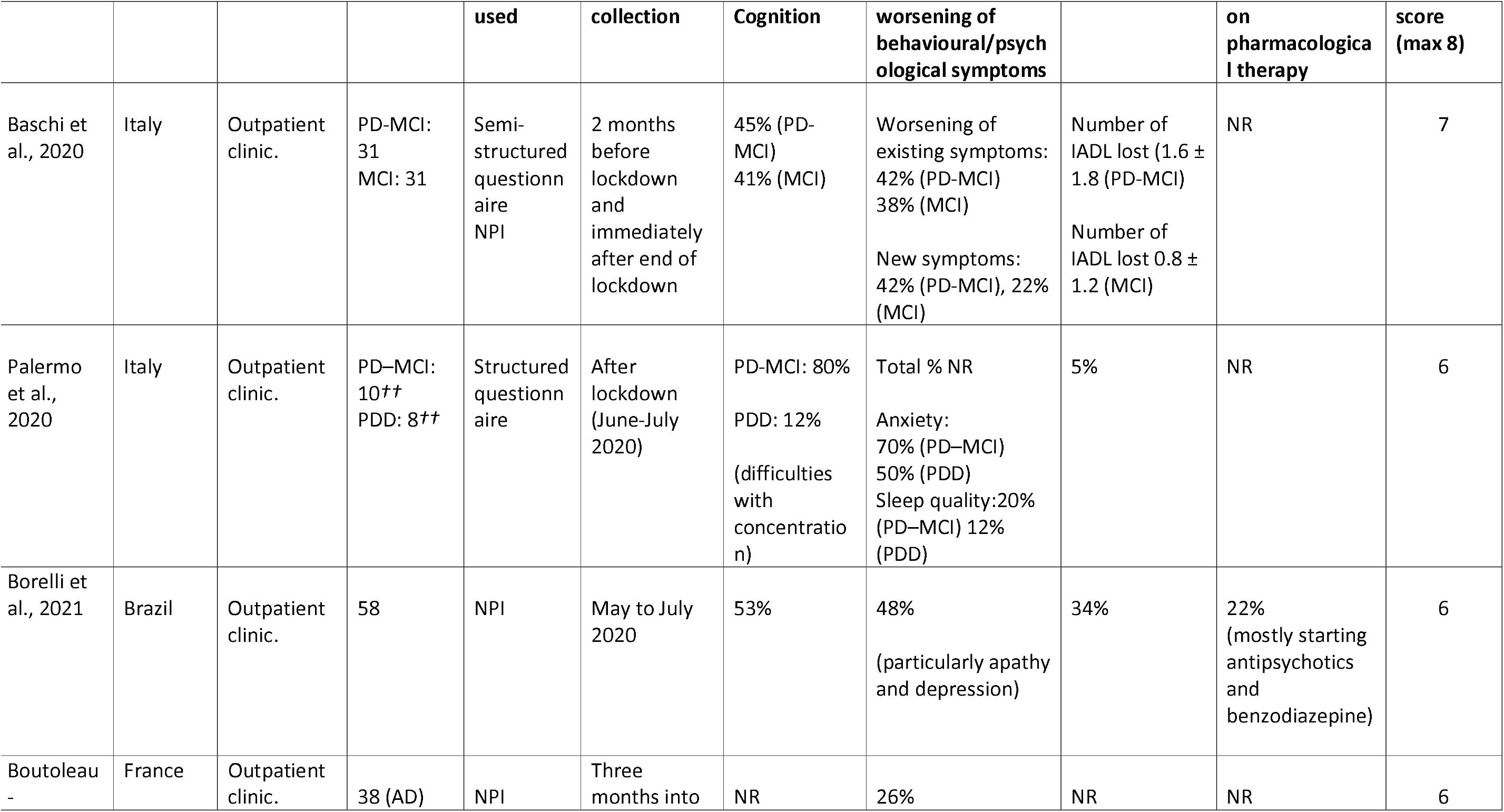
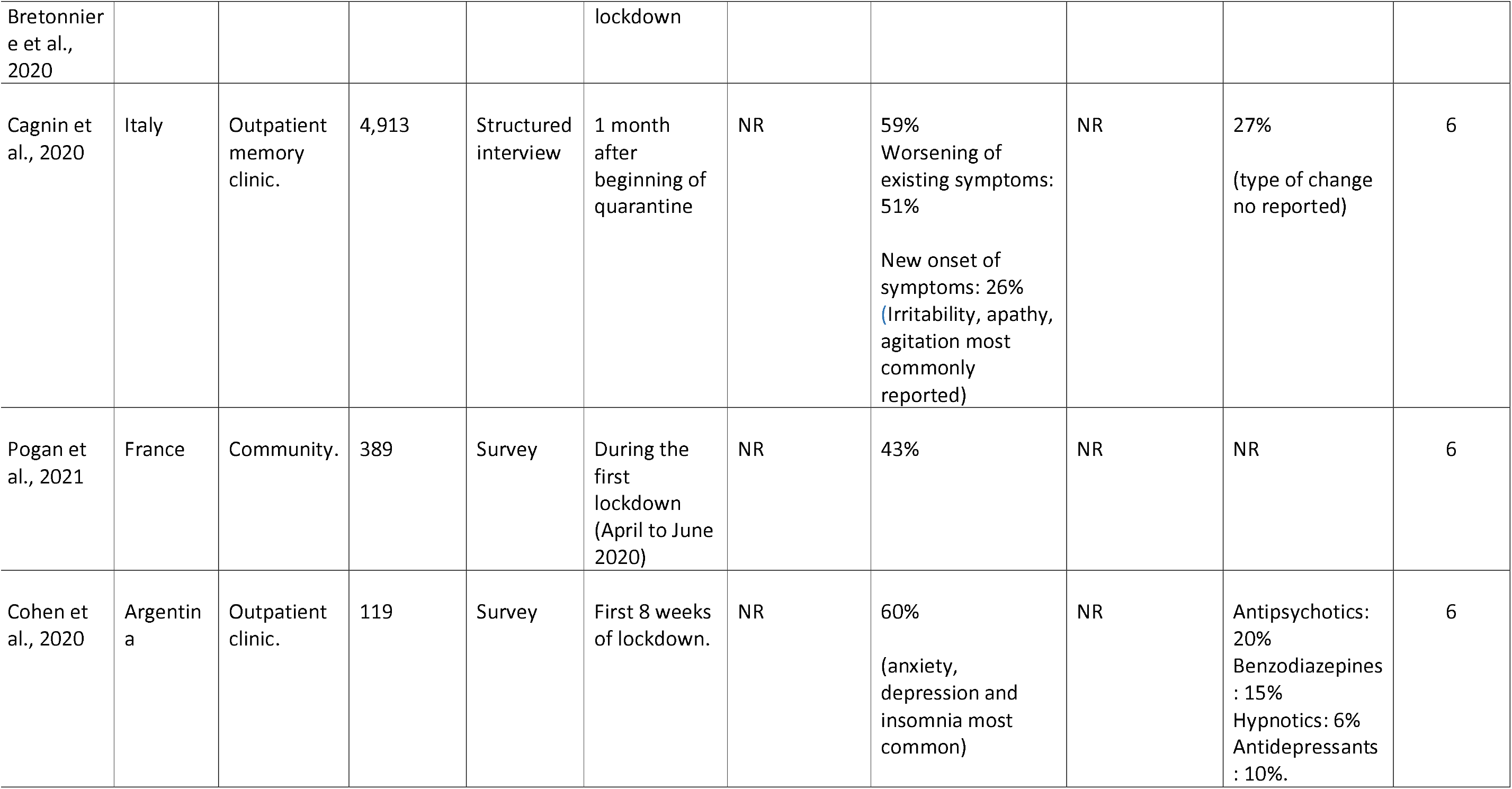
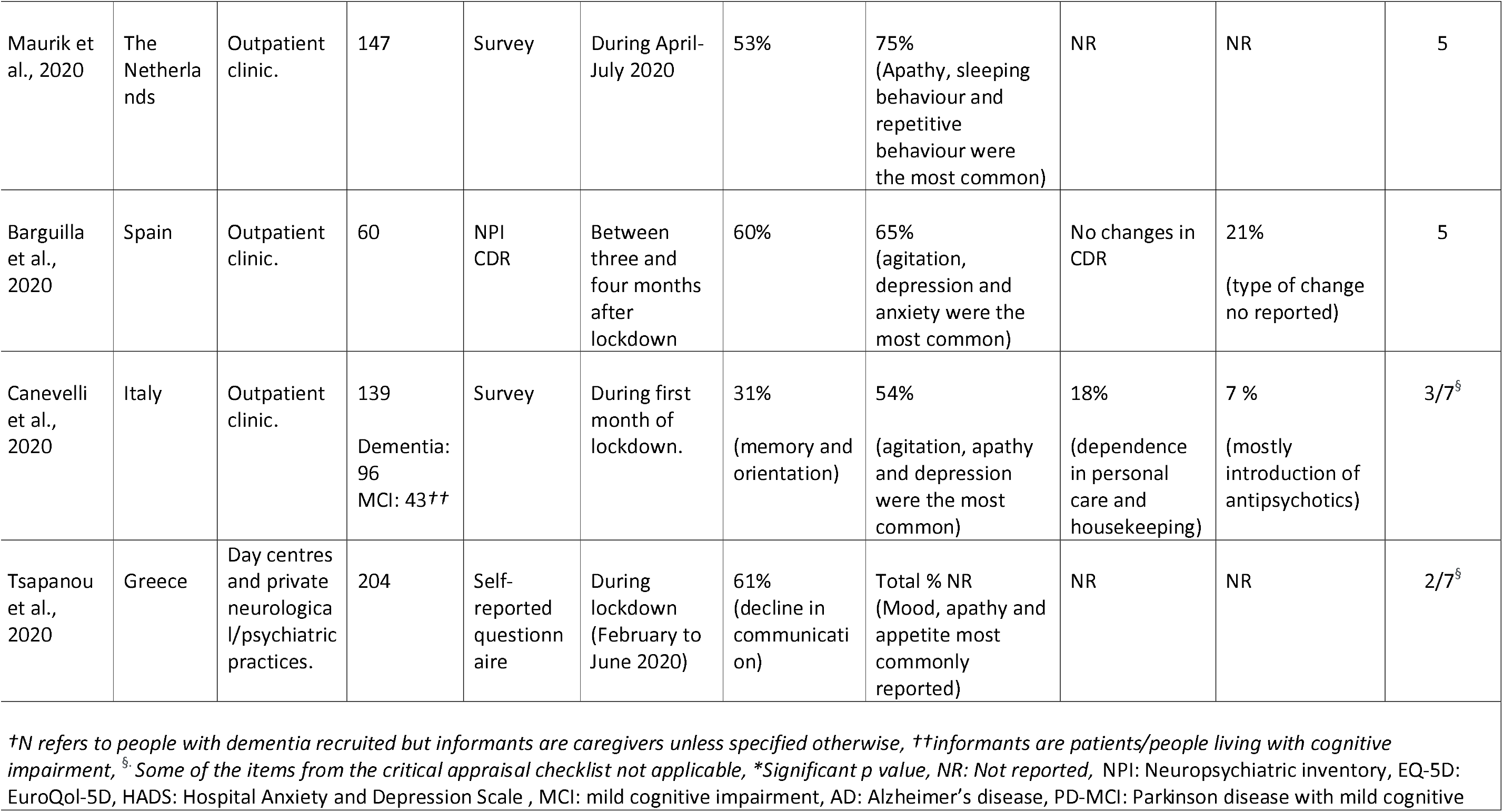
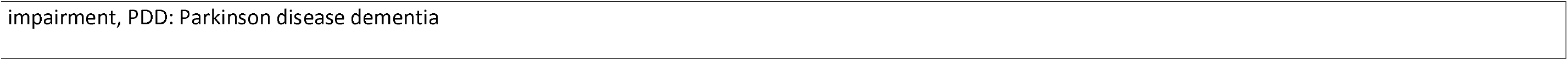
Summary of studies examining the effect of lockdown in people living with dementia in the community.

**Table 2.**
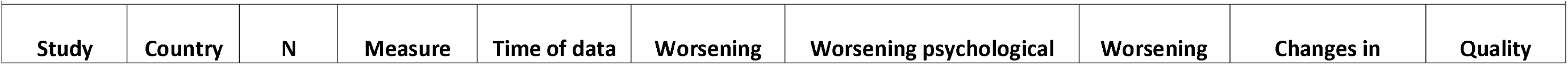
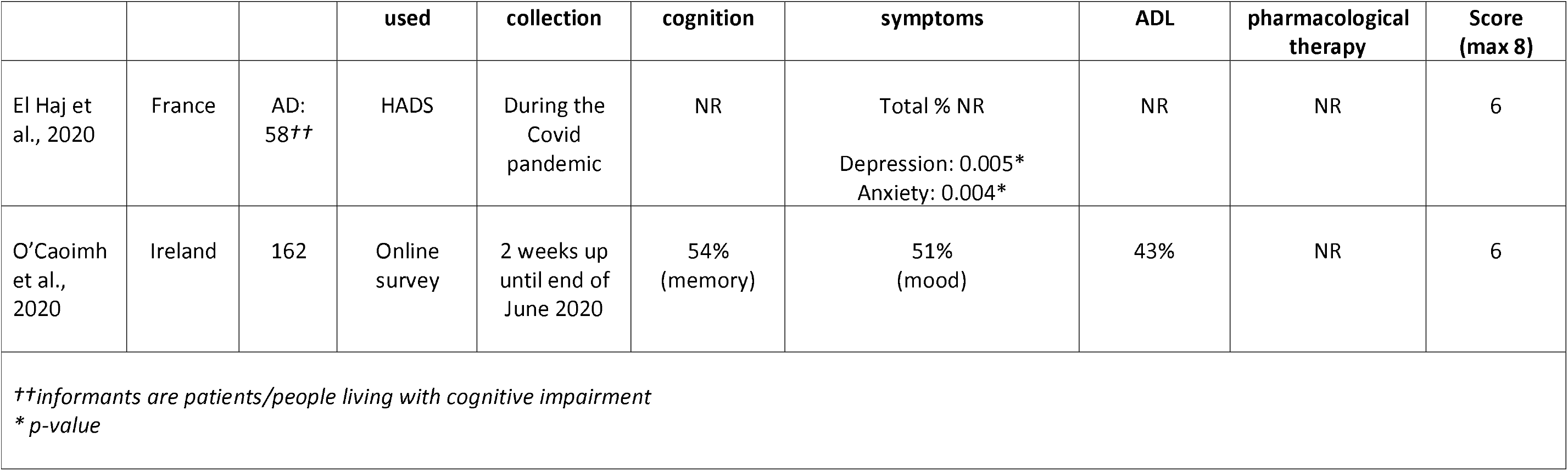
Summary of studies examining the effect of lockdown in people living with dementia in care homes.

11 studies met the criteria for good quality standard while 2 studies did not. Common quality items missing related to incorrect report of standard criteria used for diagnosis of the dementia, failure to identify and deal with confounding factors and absence of valid and reliable tools to measure outcomes.

### Studies conducted in people with dementia living in the community

#### Decline in cognition

61% (8/13) of studies reported decline in cognition. The most commonly used tool were tailored questionnaires and surveys. Reported worsening in cognition ranges 12% - 80% across studies, with 75% (6/8) of them describing declining cognition in > 50% of respondents. Concentration, memory, orientation and communication were the cognitive domains most named as affected.

#### Appearance or worsening of behavioural/psychological symptoms

92% (12/13) of studies reported worsening or new onset of behavioural and psychological symptoms, measured using questionnaires validated for the measurement of symptoms, the Neuropsychiatric Inventory (NPI) and the Hospital Anxiety and Depression Scale (HADS). Reported changes range 75%-22% in 13 patient groups. In at least 75% of the 13 groups reported more than 40% of respondents confirmed worsening or onset of new symptoms. An increase of anxiety, apathy, depression and agitation were the most common changes reported.

#### Decline in activities of daily living (ADL)

46% (6/13) studies reported changes in ADL. The tool to measure it were tailored questionnaires and Barthel Index. Reported worsening in ADL ranges 34%-5%. One study reported no changes and one significant decline in Barthel index. When the type of decline was stated, this related to the level of independence in personal care and housekeeping.

#### Increase/addition pharmacological therapy

Six studies (46%) reported adjustments to pharmacological treatment as a result of worsening neuropsychiatric symptoms during confinement, ranging from 7%-27%of respondents. When the drug was reported, this was most commonly the introduction of antipsychotics and benzodiazepines.

### Studies conducted in people with dementia living in care homes

#### Decline in cognition

One of the two studies examining the effects of lockdown in people with dementia in care homes reported changes in cognition, with 54% of residents experiencing worsening of memory, according to reports from an online survey administer to caregivers.

#### Appearance or worsening of behavioural/psychological symptoms

Mean depression and anxiety scores increased in one study and mood deteriorated in 51% of residents examined in the other.

#### Decline in ADL

One study reported worsening of independence in ADL in 43% of the residents whose caregivers responded to the survey.

#### Increase/addition pharmacological therapy

This was not reported.

## 5. Discussion

This rapid review shows evidence of worsening of cognition, neuropsychiatric symptoms and level of function for ADL in people with dementia during the Covid-19 lockdowns in Europe and Latin America at home and in care homes. They also showed an increase in prescription of antipsychotics and benzodiazepines for the psychological symptoms. The most frequently measured of these three variables neuropsychiatric symptoms, examined in 14/15 studies, followed by changes in cognition 9/15 and ADL in 7/15. Neuropsychiatric manifestations are highly disrupting and directedly associated with caregiver burden and the person with dementia quality of life, risk of care home admission, and worsening cognitive and functional outcomes (Kales et la., 2014). They can also be reported by caregiving informants without direct contact by any researcher or clinician. Hence, the practicality and interest in studying how Covid-19 has affected these specific symptoms.

The majority of the studies in this review examining behavioural and psychological symptoms reported large percentages of people living with dementia deteriorated. Nine studies reported more than 40% of participants experienced worsening or new onset symptoms (ranging 42%-75%) (Baschi et al., 2020, Palermo et al., 2020, Borelli et al., 2021, Cagnin et al., 2020, Maurik et al., 2020, Barguilla et al., 2020, Pogan et al., 2021, Canevelli et al., 2020, Cohen et al., 2020) and four others showed a similar picture of detrioration but in fewer people with dementia (ranging 12 to 38) (Cagnin et al., 2020, Boutoleau-Bretonniere et al., 2020, Palermo et al., 2020, Baschi et al., 2020). Most studies name depression, anxiety and apathy as the symptoms aggravated the most during lockdowns. Moreover, depression, along with apathy, and anxiety are strongly associated with increased caregiver burden and lower quality of life in people living with dementia (Gómez-Gallego et al., 2012; Hoe et al., 2006; Seignourel et al., 2008; Springate and Tremont, 2014) which may have contributed to situations of increasing distress for patients and families.

This rapid review also found a large proportion of people with dementia experiencing decline in their cognitive abilities due to lockdown (Baschi et al., 2020; Borelli et al., 2021 Maurik et al., 2020; Barguilla et al., 2020, Canevelli et al., 2020, Tsapanou et al., 2020). One small study with only eight people with PDD did not find many people deteriorated (Palermo et al., 2020). While people with dementia deteriorate gradually over time there is usually little difference over a few months. Data regarding the impact of Covid-19 policies on the ADL of people with dementia is scarce and more difficult to interpret due to variation in the outcomes reported (from no affectation at all to 34%) and also measurement tools (e.g. Barthel Index and questionaries).

Very little information is available about the impact of Covid-19 on people residing in care homes with only two studies addressing this (El Haj et al., 2020, O’Caoimh et al., 2020). This population has been more difficult to reach that those living in the community due to additional restrictions imposed in the social care systems (Comas-Herrera et al., 2020), which is at the same time understandable and of concern. The two studies included in this review both describing a significant worsening in psychological symptoms of residents although comparisons are impossible due to the different measures used. One of them reported a decline in cognition and ADL in around half the residents (O’Caoimh et al., 2020) which is in keeping with other emerging evidence on the topic (Leontievas et al., 2020). People with dementia in care homes are believed to have gone through the hardest version of Covid-19, both in term of mortality and deleterious effects of confinement, including decisions such as not referring to hospitals and ban on visitors which while made to protect the population can be regarded conflicting with individual human right (Suárez-González et al., 2020b). Although speculative without more data available, it is reasonable to expect, that the deleterious effect of confinement is amplified in this population compared to those living in the community because there are no relatives there, isolation has been greater and, in most homes, both activities and going out have stopped. In addition, those with dementia living in care homes tend to be older, more fragile and at more severe stages of the disease progression. It is therefore paramount that new research studies contribute to shed light on this worrying gap of evidence.

### Changes in pharmacological therapy

The one previous study on the prescription of antipsychotics for people with dementia in care homes during the pandemic have already suggested that their use increased significantly during the first months of 2020 (Howard et al 2020). Our review is in line with this notion since, in all studies that reported this variable, increases in dosage or initiation of medicating was found in up to 27% of people with dementia studied (most commonly antipsychotics and benzodiazepines). This may be indicative of generalised aggravation of dementia symptoms due to lockdown and impossibility of resorting to non-pharmacological management strategies due to the limitations imposed by anti-covid measures. It is important as both of these drugs have side effects including increased falls and for antipsychotics higher mortality.

### Limitations

Heterogeneity of tools and most importantly the choice of data reporting are the main limitation to synthesise and interpret the data from the studies identified in this review. For instance, whether a total percentage of the patient experiencing change in their symptoms was given, or whether the percentage was broken down by symptoms (with no overall figures) or whether the method used to measure the variables did not allow to extract percentages and mead and standard deviation were used instead. In addition, some measurement instruments used, such as the HADS, are not appropriate to be used in dementia. It is also fair to notice the difficulties in conducting these studies during the pandemic, in particular in care homes, that banned the entrance to external personnel and families and went through many strains to continue care throughout the pandemic.

## 6. Conclusions

The Covid-19 isolation measures have had a detrimental effect on the cognitive and neuropsychiatric symptoms of people with dementia. The long-term consequences of this accelerated decline are still unknown but most probably will imply poorer health and quality of life and reduced life expectancy for many. People with dementia face many challenges and losses as result of their condition in normal times. They have the right to receive the best care and support available to mitigate the disease’s symptoms and to promote their independence and well-being. Public infection control and prevention protocols that affect people with dementia have become a source of harm and they need, as a matter of urgency, to be redesigned under principles of compassionate fair care and non-maleficence. More specifically, this review makes the following calls for action:

1. Both family caregivers and paid carers should be prioritised for vaccines (as it is already happening in some countries) so home care is not disrupted. Of particular importance is to offer vaccination to staff at a time they can access it when working on shifts.
2. Measures to support remote working and work-life balance policies for family caregivers should be implemented and maintained until the pandemic is over and extended to more than one caregiver (some families share the caregiving tasks).
3. We now know that the risk of infection is low outdoors, and the correct use of appropriate PPE can allow safe close physical contact. This knowledge should be used to restore routines, support and therapeutic activities in the community for people with dementia.
4. Care homes in many countries are progressing in the immunisation of both residents and workers but even in the cases where this is not happening, safe visits can and should be maintained (see Storr et al., 2021, Low et al., 2021 and https://enablesafecare.org/ for a list of specific measures to enable safe human interaction in care homes).

## Data Availability

N/A

## Acknowledgements

ASG receives funding from a grant jointly funded by the Economic and Social Research Council (UK) and the National Institute for Health Research (UK) (ES/S010467/1). GL is supported by UCLH National Institute for Health Research (NIHR) Biomedical Research Centre, and by NIHR North Thames ARC as a NIHR senior investigator. JR is supported by a UK Research and Innovation ‘s Global Challenges Research Fund (UKRI GCRF) (ES/P0109381).

## Data share statement

Data sharing in not applicable to this article as no new data were created or analysed in this study.

